# Discovering methylation markers and development of a sense-antisense and dual-MGB probe PCR assay in plasma for colorectal cancer early detection

**DOI:** 10.1101/2024.07.31.24311206

**Authors:** Yanteng Zhao, Zhijie Wang, Qiuning Yu, Xin Liu, Xue Liu, Shuling Dong, Xianping Lv, Tiao Zhang, Dihan Zhou, Qiankun Yang

## Abstract

Screening for colorectal cancer (CRC) using plasma methylation is challenging due to the low abundance of cell-free DNA (cfDNA). Therefore, the development of signal amplification assays based on appropriate markers is essential to increase sensitivity. In this study, we employed an epigenome-wide approach for de novo identification differentially methylated CpGs (DMCs) common to CRC and adenoma using 17 public datasets. A sense-antisense and dual MGB probe (SADMP) assay was then developed. Subsequently, the biomarkers were validated in 712 plasma samples based on SADMP. A total of 2237 DMCs showed overlap between CRC and adenoma. Of these, 75 were hypomethylated in 30 other non-CRC cancers. Following LASSO regression, WBC validation and primer/probe design evaluation, *NTMT1* and *MAP3K14-AS1* were identified as the most informative candidate biomarkers. At preset template concentrations, the SADMP assay for *NTMT1* or *MAP3K14-AS1* could reduce the cycle threshold by 1. The *NTMT1* and *MAP3K14-AS1* dual-target SADMP assay demonstrated a sensitivity of 84.8% for CRC (stage I: 75.0%), a sensitivity of 32.0% for advanced adenomas (AA), and a specificity of 91.5% in controls. The dual-target assay showed high performance for CRC early detection in plasma, suggesting that it may serve as a promising noninvasive tool for CRC screening.

## 1. Introduction

The majority of early-stage advanced colorectal neoplasms, including colorectal cancer (CRC) and advanced adenoma (AA) are curable, particularly AA, which can be removed at the time of diagnosis by colonoscopy. Consequently, the early detection of CRC and AA represents a highly valuable endeavor. A number of tests based on cell-free DNA (cfDNA) methylation have been developed and have shown good performance for CRC detection. Epi proColon [1] is the first blood-based test utilized for the early detection of CRC and was developed based on methylated cfDNA of *Septin9*. The updated version of Epi proColon® 2.0 CE demonstrated an improved sensitivity of 74.8%-81% and specificity of 96.3%-99% in a prospective cohort study [2]. The Chinese version of Epi proColon simplified sample processing with a single reaction system in a larger volume (60 ul) instead of a 2/3 algorithm (20 ul for three runs), resulting a sensitivity of 73% and a specificity of 94.5% [3]. However, its sensitivity for detecting polyps or AA was relatively low in most studies, ranging from 8-40% [4-6]. Due to this relatively poor sensitivity, particularly for precancerous lesions, the US Preventive Services Task Force and the American Cancer Society currently do not include the test in their CRC screening guidelines. Moreover, plasma methylated *Septin9* is not a CRC specific marker but showed an ability to detect multiple cancer types, including hepatocellular carcinoma [7], gastric cancer [8], cervical cancer [9], and breast cancer [10]. Therefore, developing a sensitive and specific blood assay for CRC screening is meaningful.

Due to the disparate mechanisms by which cancer signals enter the blood and stool, it is probable that the markers are not universal, and the accuracy of CRC markers in the blood is typically low in comparison to those in stool. Consequently, an ab initio search for blood methylation markers for early CRC and AA is imperative. Furthermore, the detection of the methylation signal of cell-free tumor DNA (ctDNA) in plasma is challenging due to the presence of ctDNA fragments released by multiple organs or tissues, as well as DNA damage caused by bisulfite conversion [11]. Therefore, the identification of suitable markers and the development of effective detection techniques are essential to ensure optimal performance [12]. Classical PCR-based methylation detection techniques are often developed based on single-strand bisulfite converted DNA (BS-DNA), leaving the information of the other strand unused [13]. Furthermore, only one Taqman MGB probe is usually designed to provide fluorescent signals. Theoretically, designing primers for both sense and antisense strand DNA simultaneously or using multiple MGB probes will improve the sensitivity of a marker to detect methylation signals by enhancing fluorescent signals. Sarah Ø. Jensen et al [14] made the first attempt to design a pair of primers for both sense and antisense strands, thereby enhancing the performance of three targets for CRC detection. Additionally, the dual-strand technique was also successfully applied to methylated *HOXA9* in ovarian cancer to improve the detection sensitivity [15].

In this study, we utilized all publicly available methylation data for CRC and adenomas to identify methylation markers, with a particular focus on those that are hypermethylated in adenomas. Furthermore, in order to enhance the ability of candidate markers to detect low-abundance cfDNA methylation signals in plasma, we attempted to apply dual-strand and dual-MGB probe techniques simultaneously, which we called the sense-antisense and dual-MGB probe (SADMP) technique, to develop a novel CRC plasma test. Finally, the test performance was comprehensively assessed in our recruited training and validation cohorts.

## 2. Materials and Methods

### 2.1. Data preparation

Thirteen methylation datasets, which were GSE77954, GSE101764, GSE131013, GSE199057, GSE164811, GSE193535, GSE129364, GSE139404, GSE107352, GSE75546, GSE77965, GSE68060 and EMTAB6450 from public databases were selected based on three criteria: 1) were generated by Illumina HumanMethylation 450k BeadChip and had the raw IDAT files, 2) the sample size was greater than 10, and 3) consisted of CRC or adenoma or normal adjacent tissue (NAT). All the IDAT files were then processed using the minfi tool [16] to obtain methylation β values. They were integrated as a single dataset (n= 1165), which we defined as Phase I discovery set for candidate marker identification.

Level 3 methylation data for 31 cancer types were retrieved from The Cancer Genome Atlas (TCGA) database (https://portal.gdc.cancer.gov/). Data for 8258 primary tumor tissues corresponding to 31 cancer types and their 710 NATs were retained. The 31 cancer types included ACC (n=79), BLCA (n=412), BRCA (n=778), CESC (n=306), CHOL (n=36), CRC (n=379), DLBC (n=48), ESCA (n=183), GBM (n=137), HNSC (n=523), KICH (n=65), KIRC (n=312), KIRP (n=271), LGG (n=513), LIHC (n=374), LUAD (n=456), LUSC (n=364), MESO (n=87), OV (n=10), PAAD (n=183), PCPG (n=178), PRAD (n=495), SARC (n=257), SKCM (n=104), STAD (n=393), TGCT (n=133), THCA (n=503), THYM (n=124), UCEC (n=418), UCS (n=57), UVM (n=80). The TCGA dataset was defined as Phase II discovery dataset. DMCs were deemed eligible if they exhibited methylation levels <0.2 on other cancer types (non-CRC), >=0.55 on CRC, and <0.15 on 710 NATs.

The other three datasets, GSE48684 [17], GSE40279 [18], and GSE122126 [19], generated by the same platform of Illumina HumanMethylation BeadChip in Gene Expression Omnibus (GEO) were used as validation sets to verify the methylation status of candidate differentially methylated CpGs (DMCs). The GSE48684 cohort consisted of 105 qualified samples, of which 41 were NAT and 64 were CRC. The GSE40279 cohort comprised whole blood cell (WBC) samples collected from 656 healthy individuals. The GSE122126 cohort included three CRC and 12 normal plasma samples.

### 2.2. Sample collection

This case-control study enrolled 772 cases, including 60 tissue samples (30 CRC and 30 NATs) and 712 plasma samples from the First Affiliated Hospital of Zhengzhou University between April 2022 and June 2022. Tissue samples were obtained from the preserved formalin-fixed paraffin-embedded (FFPE) sections collected from surgical patients. The plasma cohort was randomly divided into a training set and a validation set in a 2:1 ratio. The training set comprised 474 participants, including 115 healthy blood donors, 123 non-digestive disease patients (NDD), 65 intestinal disease patients (ID), 14 polyps patients, 10 non-advanced adenoma patients (Non-AA), 16 AA, 125 CRC patients, and 6 patients with other cancers. In the validation set, there were 235 participants, including 57 healthy blood donors, 60 NDD, 30 intestinal ID, 6 polyps patients, 5 Non-AA, 9 AA, 66 CRC patients, and 2 patients with other cancers (**Supplemental Table 1**). This study is a sub-project of the Clinical Study of Pan-cancer DNA Methylation Test in plasma (Clinical Trials ID: NCT05685524). Patients with polyps, adenomas, CRC and other cancers were confirmed by histopathological examinations. The included CRC patients were required to meet the following criteria: 1) did not receive any radiotherapy, chemotherapy or surgery, 2) ages older than 18. All CRC patients were classified as I, II, III, and IV stages according to the American Joint Cancer Committee (AJCC) staging system.

### 2.3. Differential methylation analysis

We performed differential methylation analysis using the rank-sum test on three groups in the discovery set: cancer vs. normal (NAT), adenoma vs. normal, and cancer vs. adenoma. DMCs were defined as significant if they meet two criteria: an FDR < 0.05 and a fold change ranking in the top 1% of all probes. The DMCs were then classified as hyper- or hypo- DMCs based on whether they exhibited high or low methylation levels in the tumor or adenoma samples compared to the normal samples. LASSO regression was implemented in the R package ‘glmnet’ to reduce the number of features. LASSO regression was repeated 100 times, and the frequencies of probes with non-zero coefficients in the regressions were counted.

### 2.4. Tissue and plasma DNA extraction

Genomic DNA from tissue samples and plasma cfDNA was isolated using the FFPE DNA rapid extraction kit (TianGen, Beijing) and the plasma/serum cfDNA extraction kit (Ammunition Life-tech, Wuhan), respectively, according to the protocols. Briefly, cfDNA extraction was divided into two steps. First, approximately 10 ml of blood was drawn from participants at room temperature and then centrifuged twice with 1300g and 14000g for 10 min at 4°C within 2 hours. Then, about 2 ml of plasma was retained for cfDNA purification. Purified cfDNA was eluted in 45 ul TE buffer and then treated with bisulfite using a DNA bisulfite modification kit (Ammunition Life-tech, Wuhan) in accordance with the instructions described in the aforementioned study [20], or stored at -80 °C until required.

### 2.5. Sanger sequencing

Sanger sequencing (ABI 3730XL, GENEWIZ, Suzhou) was performed for the target regions of *NTMT1* and *MAP3K14-AS1* after bisulfite treatment to confirm their methylation status on 30 CRC tissue samples and 30 NATs. The sequencing primers are shown in **Supplemental Table 2**. Following bisulfite treatment, cytosine in methylated CpG sites remained cytosine, while unmethylated cytosine was converted to thymine. Sanger sequencing results thus allowed for the direct determination of the methylation status of target regions.

### 2.6. Developing the SADMP and methylation-specific PCR (MSP) technique

The location and sequences of MSP primers/ probes are displayed in **Supplemental Table 3**. For *NTMT1*, two pairs of methylation primers targeted the sense and antisense strands, and their corresponding MGB probes were designed. For *MAP3K14-AS1*, the MGB-probe 1 and MGB-probe 2 were designed according to the antisense strand sequence and the reverse complementary sequence of the antisense strand.

Gradient dilutions of synthetic plasmids or plasmid mixtures are employed as templates for the determination of test parameters. Specifically, standard curve experiments were carried out to assess the amplification efficiency of each pair of primers. The experiments consisted of five ten-fold dilutions of fully methylated plasmid DNA templates with five replicates at each dilution (1, 10, 100, 1000, and 10000 copies per run). Primer tolerance experiments were designed to assess the specific amplification ability of methylated primers against methylated templates. Eight concentration dilutions of fully methylated DNA templates (0, 1, 5, 10, 50, 100, 200, and 400 copies/μL) mixed with a high concentration of unmethylated plasmid (10^7^ copies/μL) were prepared to evaluate the ability of methylated primers selectively amplifying methylated templates under the background of unmethylated templates. Each concentration experiment was repeated five times.

The amplification system of MSP was shown in **Supplemental Table S4**. The test employed a triplex PCR with a FAM channel for the internal reference gene *ACTB*, a ROX channel for *NTMT1* (dual-strands), and VIC channel for *MAP3K14-AS1* (dual-MGB probes). The PCR procedure was pre-denaturation at 95°C for 5 min (step 1), denaturation at 95°C for 15s (step 2), annealing at 60°C for 30s (step 3), repeating step 2 and 3 forty-five times in plasmid samples and fifity times in plasma samples on the 7500 Fast Real-Time PCR System (Applied Biosystems, USA), the cycle threshold (Ct) was assigned to 45 or 50 for the target without amplification.

### 2.7. Statistical analysis

The data processing and analysis in this study were conducted using R software (version 4.1.0). The ‘glm’ function was employed to fit the logistic regression model with a parameter of ‘family=binomial’. Receiver operating characteristic (ROC) curve analysis was performed using the ‘pROC’ package, and the area under the ROC curve (AUC) was subsequently calculated to assess the test classification performance. The optimal sensitivity and specificity of the test were estimated when Youden’s index reached its maximal value. Sensitivity and specificity were calculated using the following formulas:

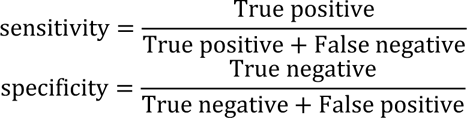

And the Youden index = sensitivity+specificity-1.

The rank-sum test and Kruskal test were employed for the comparisons between two groups and the comparisons between multiple groups, respectively. The Chi-square test was utilized for the comparisons between categorical variables. The Other statistical methods used in this study were described in the corresponding results.

## 3. Results

### 3.1. Study design and participant characteristics

The study is divided into four steps, as illustrated in **Figure 1**. In the initial step, candidate markers were identified in a set of 1165 samples from 13 datasets. Of these, 452 were normal, 168 were adenomas, and 545 were CRC. These markers were then narrowed down using data from 31 cancer types in the TCGA dataset, DMCs that were hypermethylated in cancer other than CRC were filtered out. The available DMCs were further validated in three independent datesets. In the second step, the methylation status of candidate CpGs was confirmed by Sanger sequencing using 30 CRC tissues and 30 NATs. In the third step, the SADMP technique and MSP system were established. This involved designing appropriate primers, optimizing the qPCR amplification system, and evaluating the technical parameters of the assay. In the last step, the developed assay was tested on training and validation sets of plasma. Its performance was assessed by estimating indicators such as sensitivity, specificity, and AUC.

**Figure 1.**
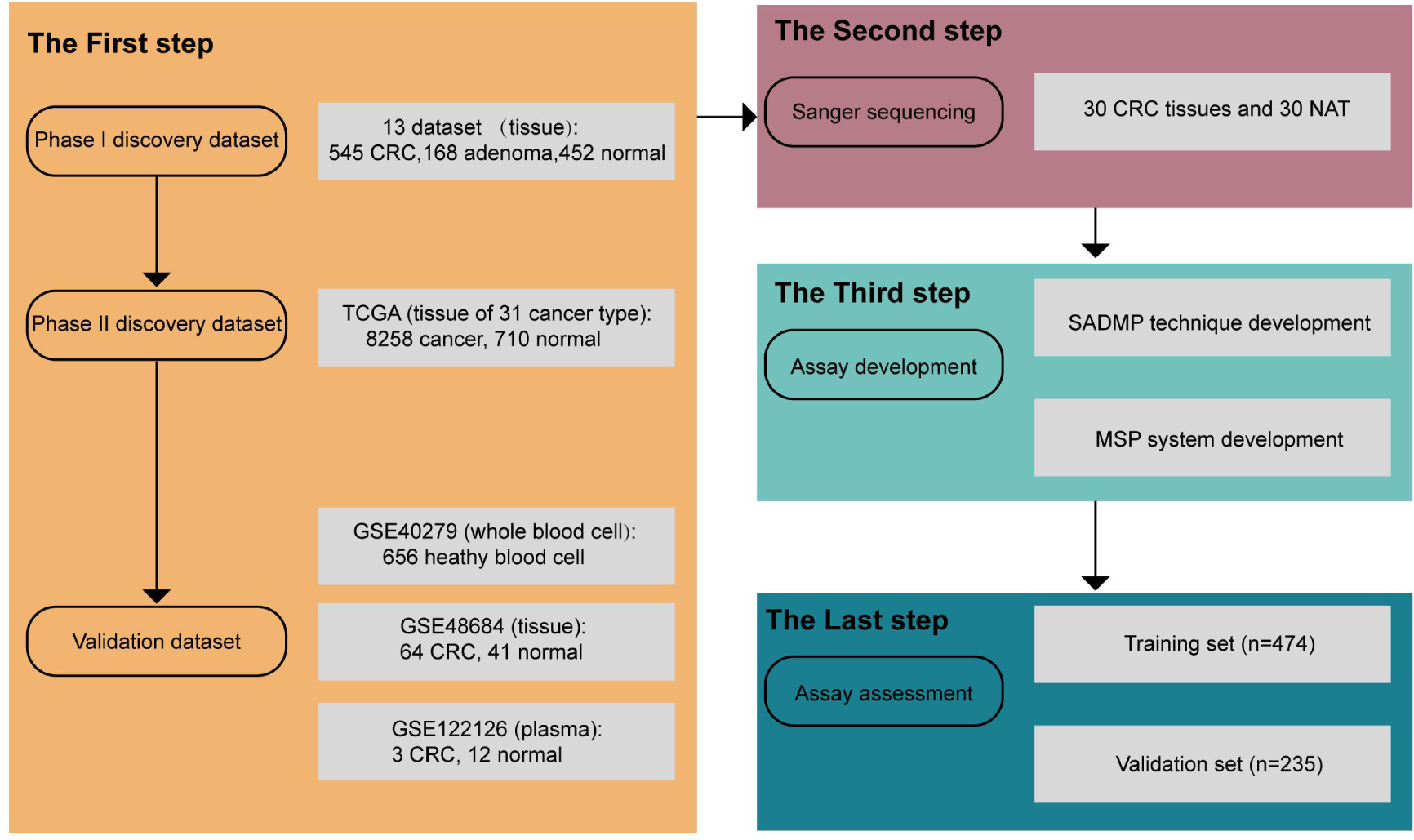
The flowchart of this study. The four steps were illustrated by different colored blocks.

### 3.2. Landscape of the methylation patterns of the 13 discovery datasets

The methylation levels of adenoma and cancer samples were found to be lower than normal, as shown in **Figure 2A**. The methylation density curves for the three groups displayed bimodal distributions with hyper- and hypomethylation peaks, respectively (**Figure 2B**). The hyper-methylation peaks of adenoma and tumor samples were lower than those of normal samples, indicating higher methylation levels in normal samples.

**Figure 2.**
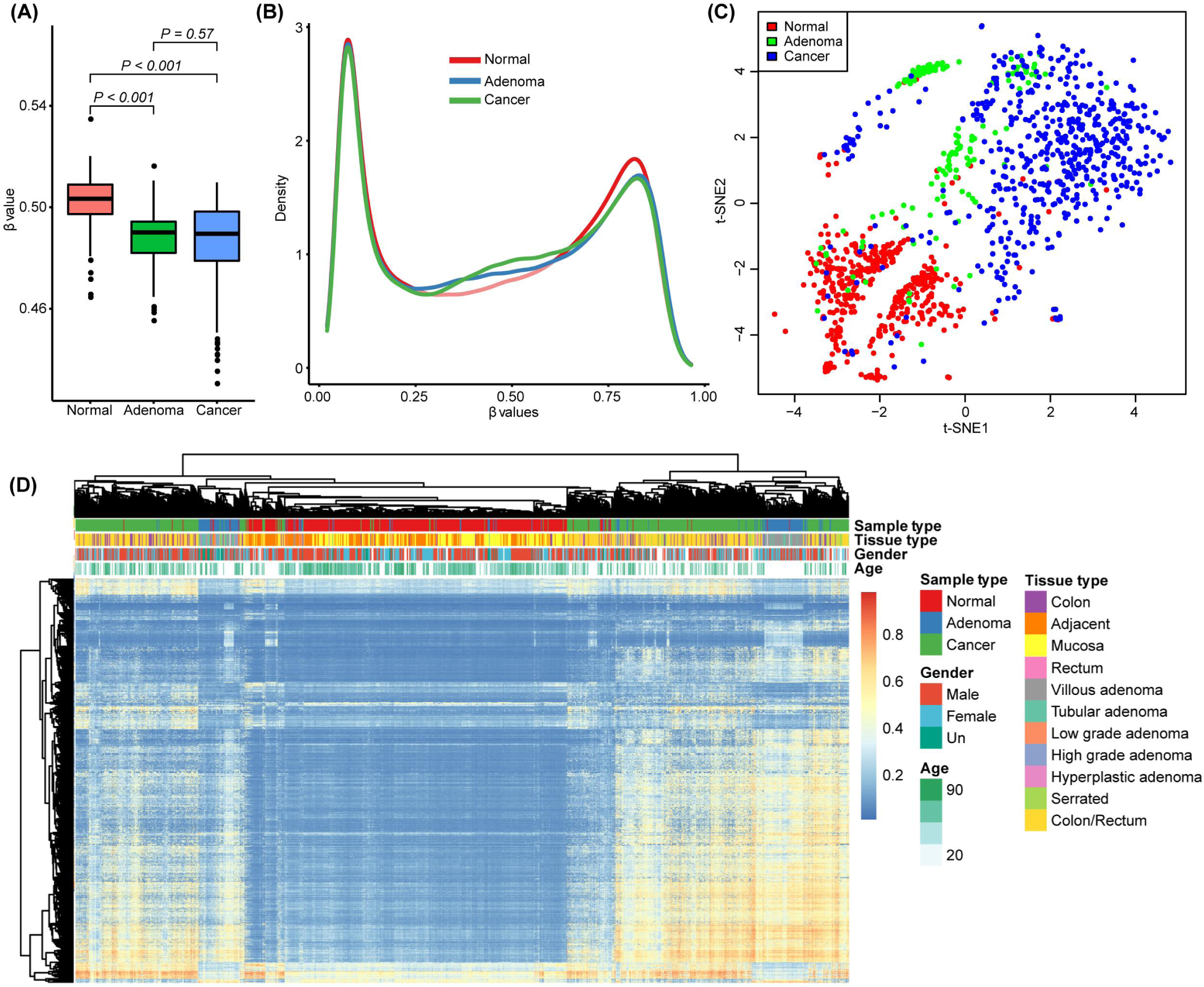
Landscape of the methylation patterns of the phase I discovery set. (**A**) boxplot showing the overall methylation levels of normal, adenoma and cancer samples. The average β value of all probes for each sample was calculated as the sample overall methylation level. P-values were estimated by Kruskal test. (**B**) Density curves of probe methylation levels in normal, adenoma and cancer samples. (**C**) t-SNE visualizing normal, adenoma and cancer samples in the discovery set. (**D**) Heatmap showing the most variable probes between normal, adenoma and cancer samples.

We employed t-SEN to analyze and visualize the structure of the discovery set and identified significant differences between normal and cancer samples. However, adenomas exhibited overlap with both normal and cancer samples (**Figure 2C**). We then selected the top 1% of most variable probes to cluster the discovery set using the K-means algorithm. The results showed that both normal and cancer samples clustered together, while adenomas were separated into two subgroups with a remarkable difference. They showed both high (Methy-H) and low (Methy-L) methylation status and were closer to cancer and normal samples, respectively (**Figure 2D**).

Further analysis revealed that tubular adenomas constituted the largest proportion of Methy-L adenomas (23/57 or 40.35%), while villous adenomas constituted the largest proportion of Methy-H adenomas (47/73 or 64.38%). To eliminate any potential bias between the datasets, we conducted Fisher’s test separately for the Methy-H and Methy-L adenomas, with the datasets serving as the stratification factor. However, we did not identify any significant differences (P > 0.05).

### 3.3. Identification of candidate DMCs

First, we conducted a comparative analysis of the DMCs between cancer, adenoma, and normal samples using the 13 discovery datasets. The three comparisons (cancer vs. normal, adenoma vs. normal, and cancer vs. adenoma) yielded 3000, 3051, and 1545 DMCs, respectively. Notably, these DMCs were predominantly hyper-DMCs (**Figure 3A**). Further investigations revealed that the majority of DMCs were distributed in upstream regions of genes (5’UTR, TSS1500, TSS500, and 1st Exon), except for cancer vs. adenoma where DMCs were mainly located in gene bodies (**Figure 3B**). We observed an extremely high proportion of overlapping DMCs between cancer vs. normal and adenoma vs. normal, which was significantly higher than cancer vs. adenoma (**Figure 3C**). To identify the most appropriate DMCs, we focused on those that were overlapped between cancer vs. normal and adenoma vs. normal (2237 probes) and evaluated their methylation levels on 31 cancer types of TCGA. This yielded 75 accessible probes (**Figure 3D**). LASSO regression was employed to reduce the number of DMCs, with eight presented in 100 replicates with non-zero coefficients (**Figure 3E**).

**Figure 3.**
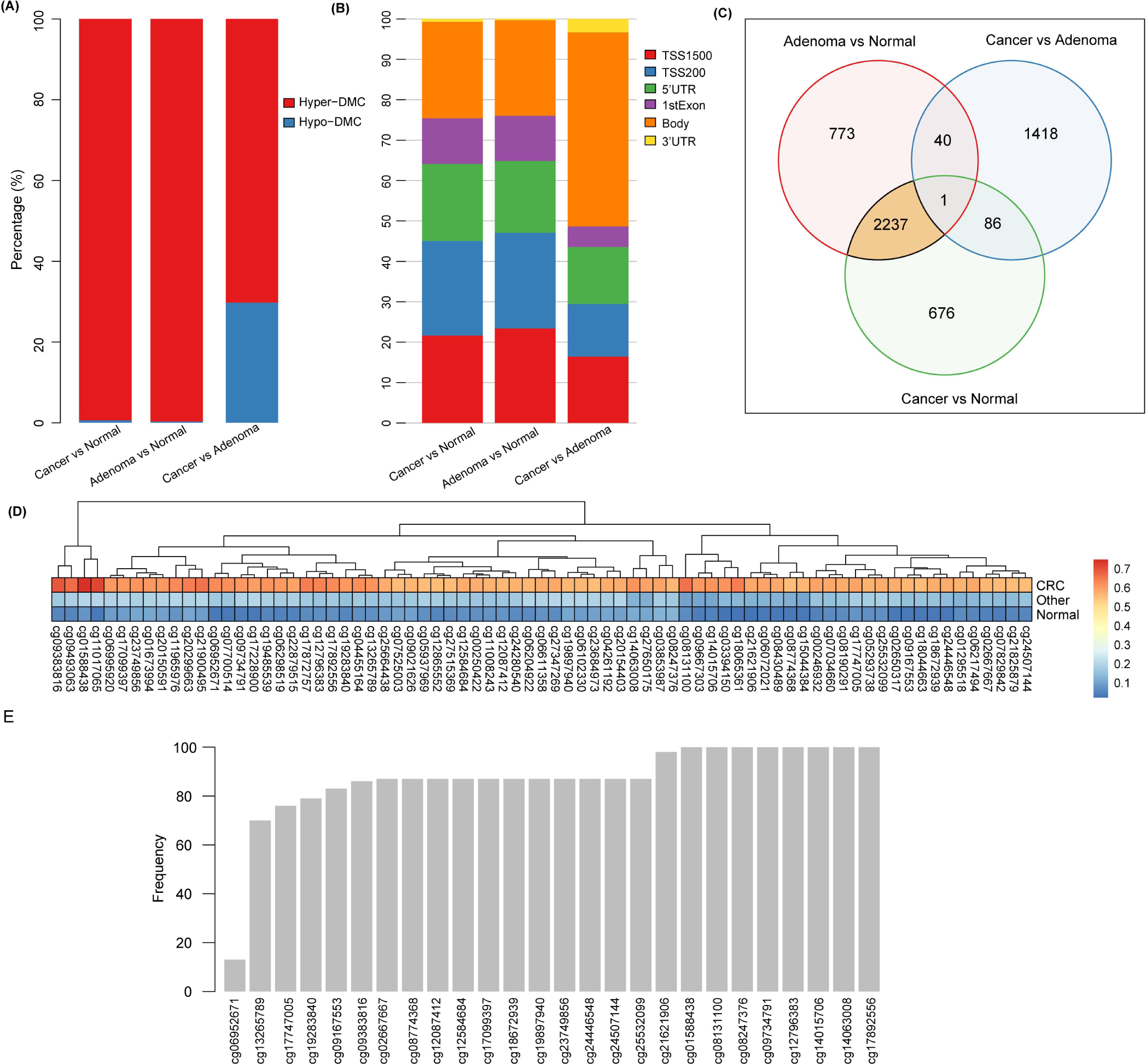
Identification of candidate markers. (**A**) Percentage of hyper-DMC and hypo-DMC between the three comparisons. (**B**) Percentage of DMC at different genomic regions between the three comparisons. (**C**) Venn diagram showing DMCs between the three comparisons. (**D**) Methylation values of 75 probes meeting the criteria on TCGA 31 cancer types. The “other” refers to 30 non-CRC cancer samples. “Normal” refers to 710 NATs. (**E**) Frequency of probes with non-zero coefficient in 100 LASSO regressions.

### 3.4. The methylation levels of NTMT1 and MAP3K14-AS1 in validation sets

The methylation levels of the eight probes were then analyzed in 656 WBC samples, and seven DMCs exhibited relatively low methylation levels (less than 0.1) with the exception of cg17892556 (**Figure 4A**). Therefore, the seven DMCs are recognized as the most promising markers.

**Figure 4.**
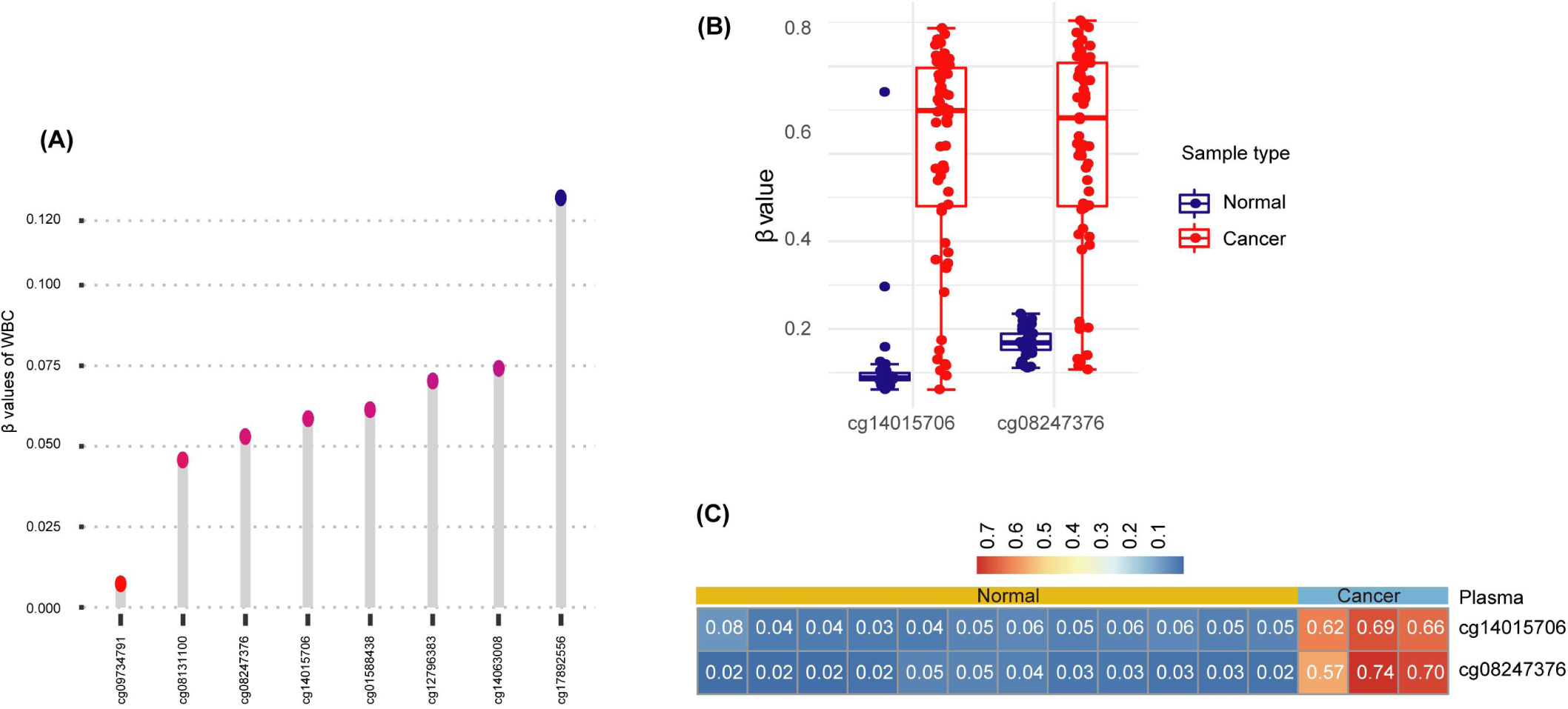
Methylation profiles of the two candidate probes in different datasets. (**A**) The methylation profiles of cg14015706 and cg08247376 in 656 healthy WBC in GSE40279. (**B**) The methylation β values of cg14015706 and cg08247376 between normal and cancer samples in GSE48684 dataset. (**C**) The cfDNA methylation profiles of cg14015706 and cg08247376 in normal and cancer plasma samples from GSE122126. Numbers in the heatmap indicated the methylation β values.

Two specific DNA methylation sites, cg14015706 and cg08247376, were selected for further analysis as they were suitable for primer/probe design. These sites are located in the first exon of *NTMT1* and 200 bp upstream of the *MAP3K14-AS1* transcription start site, respectively. The GSE48684 dataset revealed significantly higher methylation levels for both probes in cancer samples than in normal samples (**Figure 4B**). Furthermore, the two probes demonstrated hypermethylation in CRC plasma and hypomethylation in healthy plasma (**Figure 4C**), indicating a high consistency of the methylation status between tissue and plasma.

### 3.5. The SADMP technique

The Sanger sequencing of the target regions revealed that the CpGs sites among the amplicons were more frequently methylated in CRC than NATs, which provided the foundation for the development of SADMP technique. For *NTMT1*, two pairs of methylation primers targeted the sense and antisense strands, and their corresponding MGB probes were designed (**Figure 5A**). For *MAP3K14-AS1*, the MGB-probe1 and MGB-porbe2 were designed according to the antisense strand sequence and the reverse complementary sequence of antisense strand (**Figure 5F**). The estimated amplification efficiencies of *NTMT1* and *MAP3K14-AS1* were 98.14% and 110.08%, respectively (**Figure 5B** and **5G**). The sense and anti-sense strands were transformed into completely different DNA sequences after bisulfite treatment and both could be used as PCR templates. It can be postulated that the copy numbers detected by any single pair of primers or single MGB-probe assay should be half of the diluted copy numbers calculated as the sum of sense and anti-sense strand templates, and that the SADMP assay should be equal to the diluted copy numbers. For *NTMT1*, the detected copy numbers by sense and antisense strand assays were slightly less than half of the theoretical copy numbers with slopes of 0.39 and 0.31, respectively. The detected copy numbers by dual-strand assay were approximately 0.73 times of theoretical copy numbers, which was approximately two-fold compared to any single-strand assay (**Figure 5C**), as expected. Meanwhile, Ct value of dual-strand assay was almost one cycle earlier than that of any single-strand assay (average △Ct=1.21, **Figure 5D**), which was consistent with the theoretical △Ct value (should be one). The *MAP3K14-AS1* SADMP showed similar results, with the number of copies detected was doubled compared to the single probe assay (**Figure 5H**), and the Ct values were shifted forward by 0.99 cycles (**Figure 5I**).

**Figure 5.**
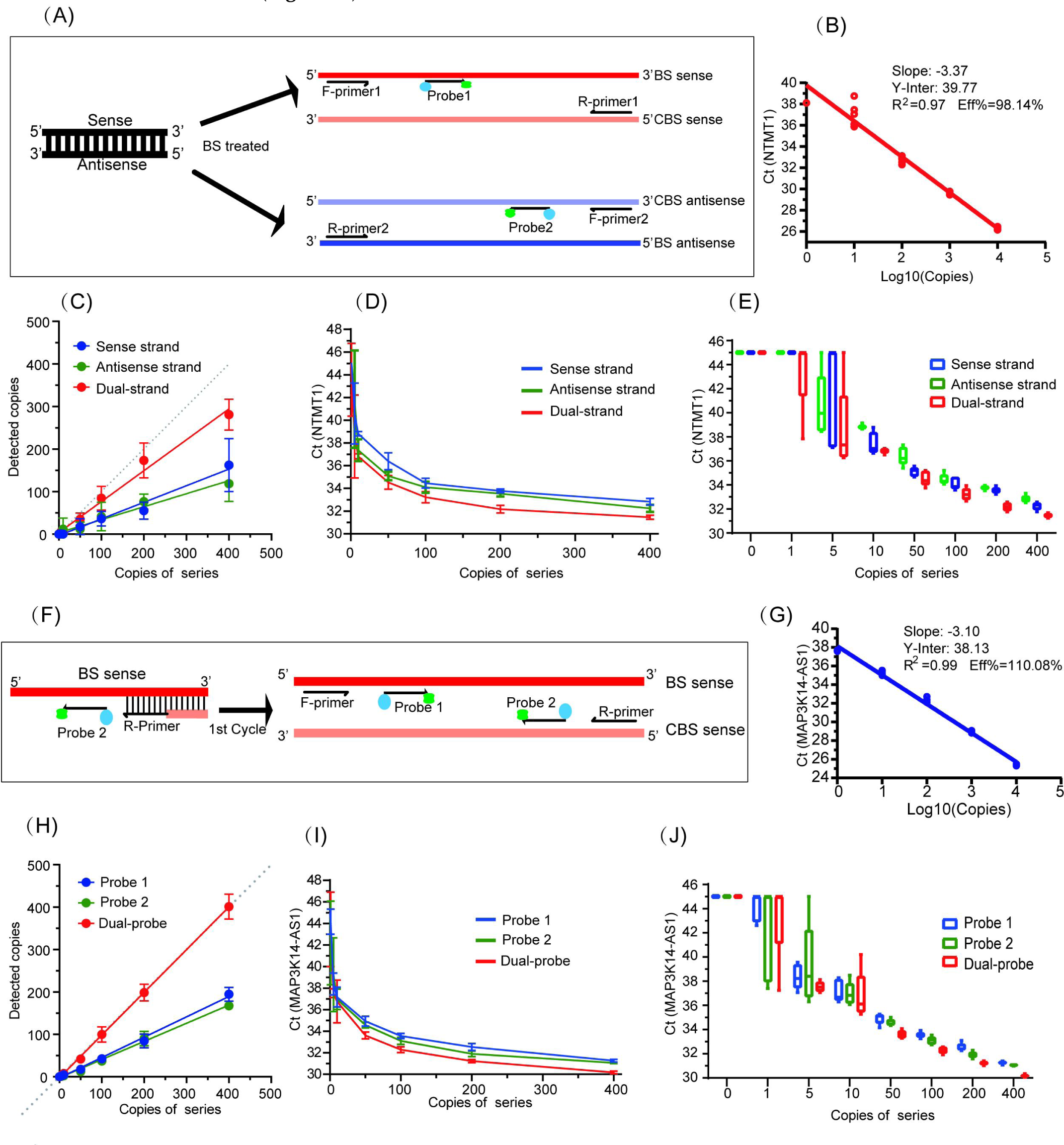
The development and validation of SADMP. (**A**) SADMP strategy for *NTMT1*. Two pairs of primers were designed according to the sense and antisense BS-strand sequences. CBS indicated the complementary sequence of sense and antisense BS-strands. (**B**) The standard amplification curve of *NTMT1*. (**C**) The agreement between detected copies by SADMP and single-strand assays and diluted template DNA copies (theoretical copies) of *NTMT1*. The solid lines were fitted by a simple linear model. (**D**) and (**E**) The detected Ct values of SADMP *NTMT1* and single-strand *NTMT1* assays for different diluted concentrations. (**F**) SADMP strategy for *NTMT1*. Two MGB probes were designed. Probe 1 is located downstream of the forward primer, targeting the BS-strand template. Probe 2 is located downstream of the reverse primer, targeting the complementary sequence of BS-strand. (**G**) The standard amplification curve of *MAP3K14-AS1*. (**H**) The agreement between detected copies by SADMP and single-probe assays and diluted template DNA copies (theoretical copies) of *MAP3K14-AS1*. The solid lines were fitted by a simple linear model. (**I**) and (**J**) The detected Ct values of SADMP *MAP3K14-AS1* and single-probe *MAP3K14-AS1* assays for different diluted concentrations.

Additionally, in the serially diluted experiment, no amplification cures for both targets were observed when a high proportion of unmethylated DNA was used as templates (the methylated DNA copies were 0) (**Figure 5E** and **5G**). These results suggested that the developed assays were explicitly targeted for methylated DNAs even at the high background of unmethylated DNAs. We also found that the *NTMT1* SADMP assay was able to robustly detect methylated DNAs at a concentration of 10 copies/μL (**Figure 5E**), while it was 5 copies/μL for the *MAP3K14-AS1* SADMP (**Figure 5G**).

### 3.6. Validation and evaluation of biomarkers using MSP in plasma samples

In the plasma sample set, the Ct values of *ACTB* exhibited a slight decrease in trend from the healthy control groups to the cancer group (p < 0.0001) (**Figure 6A**). Ct values of *NTMT1* and *MAP3K14-AS1* were much lower in CRC samples and AA samples than other control groups (**Figure 6B** and **6C**).

**Figure 6.**
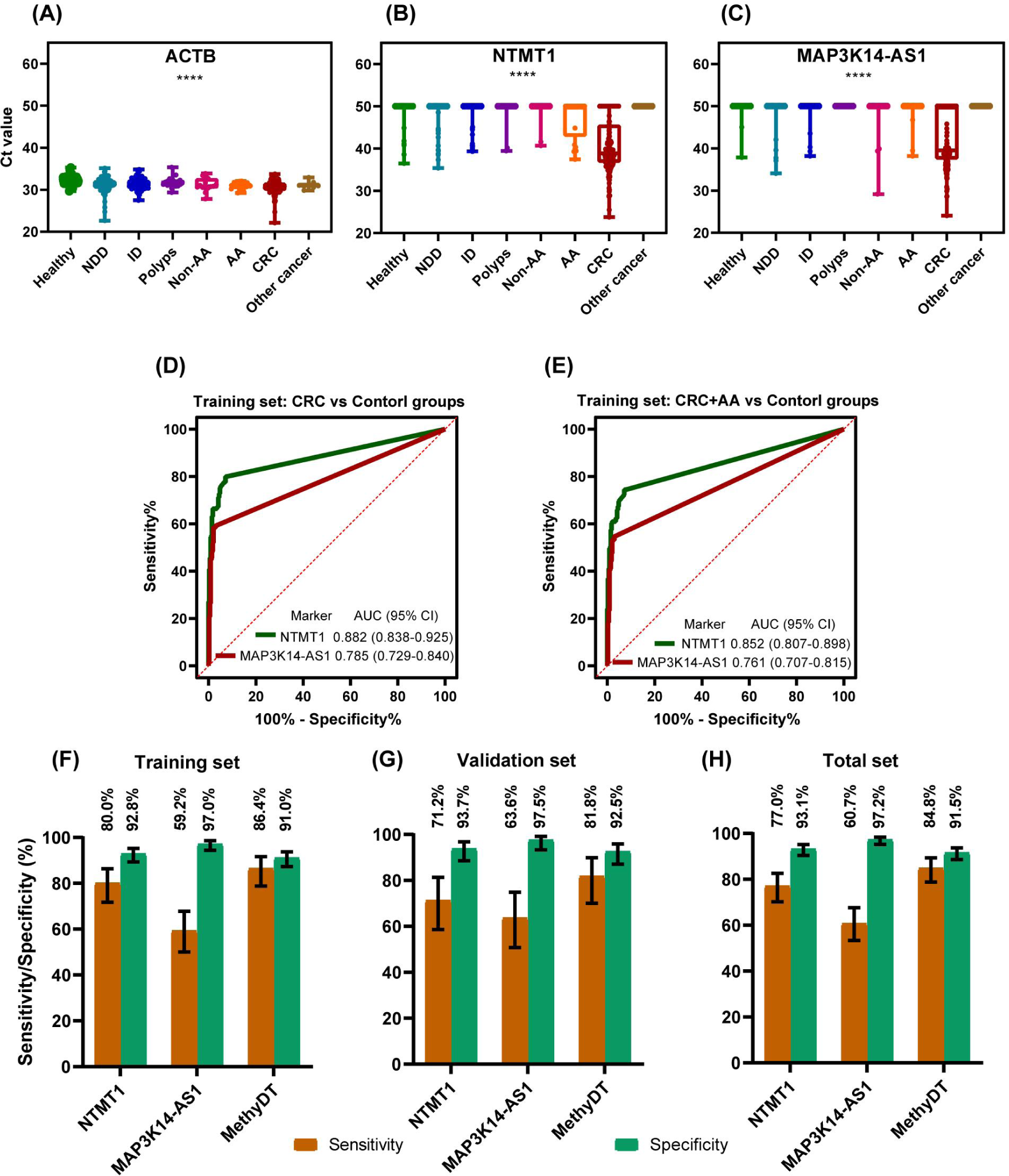
dual-target assessment and validation in plasma samples. (**A**-**C**) Ct values of *ACTB* (A), *NTMT1* (B) and *MAP3K14-AS1* (C) in various clinical groups. NDD: Non-digestive disease, ID: Intestinal disease, AA: advanced adenoma. The eight groups were compared by Kruskal-Wallis test. P<0.05 is considered significant. **** is p<0.0001. (**D**-**E**) ROC curves of two biomarkers in CRC vs Control groups (D) and CRC+AA vs Control groups (E) in training set. (F-G) Sensitivity and specificity of individual biomarker and dual-target in training set (F), validation set (G) and total set (H) using 1/2 algorithm. Error bars represent 95% CI.

The Ct values could discriminate the CRC and AA samples from healthy samples very well. Furthermore, the Ct values-based result determination exhibited favorable outcomes in a previous study [21,22]. Consequently, the Ct values were utilized as the result analysis indicators. In order to determine the algorithm and cutoff, ROC curve analysis was conducted in the training set. In the CRC vs control groups, the AUC values for *NTMT1* and *MAP3K14-AS1* were 0.882 (95% CI: 0.838-0.925) and 0.785 (95% CI: 0.729-0.840), respectively (**Figure 6D**). In the CRC+AA vs control groups, the AUC values for *NTMT1* and *MAP3K14-AS1* were 0.852 (95% CI: 0.807-0.898) and 0.761 (95% CI: 0.707-0.815), respectively (**Figure 6E**).

As AA was precancerous lesions, it was also benefit patients from detecting it. So we use the CRC+AA vs control groups of training set for further analysis. The performance of the combination of the two biomarker were evaluated by two strategies (**Table 1**). Strategy 1 involved the construction of a logistic regression model, with an estimated AUC value was 0.881 (95% CI: 0.840-0.922), the optimal sensitivity for CRC and specificity of 86.4% and 91.0%, respectively (**Table 1**). Strategy 2 was the 1/2 algorithm, which determined a positive measurement when the Ct of any single marker was less than its corresponding threshold. The cutoff was determined as Ct value corresponding to the maximum Youden index in the training dataset using CRC+ AA vs control data. The cutoff values were 48.2 and 48.4 for *NTMT1* and *MAP3K14-AS1*, respectively. At these cutoff, the two target sensitivities were 80.0% and 59.2%, with specificities of 92.8% and 97.0%, respectively (**Figure 6F**). The sensitivity and specificity of the combination of the two biomarkers were 86.4% and 91.0% respectively. Notably, strategy 2 yielded an identical sensitivity and specificity to strategy 1. Given that strategy 2 is considerably more straightforward for physicians to interpret test results in clinical practice, we have adopted the 1/2 algorithm as the combination algorithm for the dual-target test.

**Table 1.** Diagnostic performance of single biomarker and marker combination in training set.

| Biomarker & combination | AUC (95% CI) | Sensitivity (CRC) | Sensitivity (CRC+AA) | Specificity | Cutoff | Combination method |
| --- | --- | --- | --- | --- | --- | --- |
| *NTMT1* | 0.852 (0.807-0.898) | 80.0% | 74.5% | 92.8% | 48.2 | / |
| *MAP3K14-AS1* | 0.761 (0.707-0.815) | 59.2% | 54.6% | 97.0% | 48.4 | / |
| *NTMT1* & *MAP3K14-AS1* | 0.881 (0.840-0.922) | 86.4% | 80.8% | 91.0% | 0.1491 | Logistic regression |
| *NTMT1* & *MAP3K14-AS1* | / | 86.4% | 80.8% | 91.0% | 48.2, 48.4 | 1/2 algorithm |

After the algorithm and cutoff was set, the validation set and all sample set were analyzed. The validation set had a 81.8% sensitivity and 92.5% specificity (**Figure 6G**) and the all sample set had a 84.8% sensitivity and 91.5% specificity (**Figure 6H**). The dual-target test had a higher sensitivity than that of any single biomarker and a slightly lower specificity in the three data sets (**Figure 6F-H**). It indicated that the combination of the two biomarkers were better than single biomarker.

### 3.7. Performance in subgroups of plasma samples

The performance of the dual-target test was evaluated in subgroups of all plasma samples. The test demonstrated a detection rate of 26.7% in non-AA samples and a detection rate of 32.0% in AA samples (**Figure 7A**). It exhibited sensitivities of 75.0%, 81.2% and 90.3% in stages I, II and III-IV of CRC samples, respectively (**Figure 7B**). The test exhibited specificities of 95.9%, 92.3%, 84.2% and 90.9% in healthy, NDD, ID and polyps samples, respectively (**Figure 7C**). The positive predictive value (PPV), negative predictive value (NPV) and accuracy for the dual-target test were 79.4%, 94.0% and 89.6%, respectively (**Figure 7D**).

**Figure 7.**
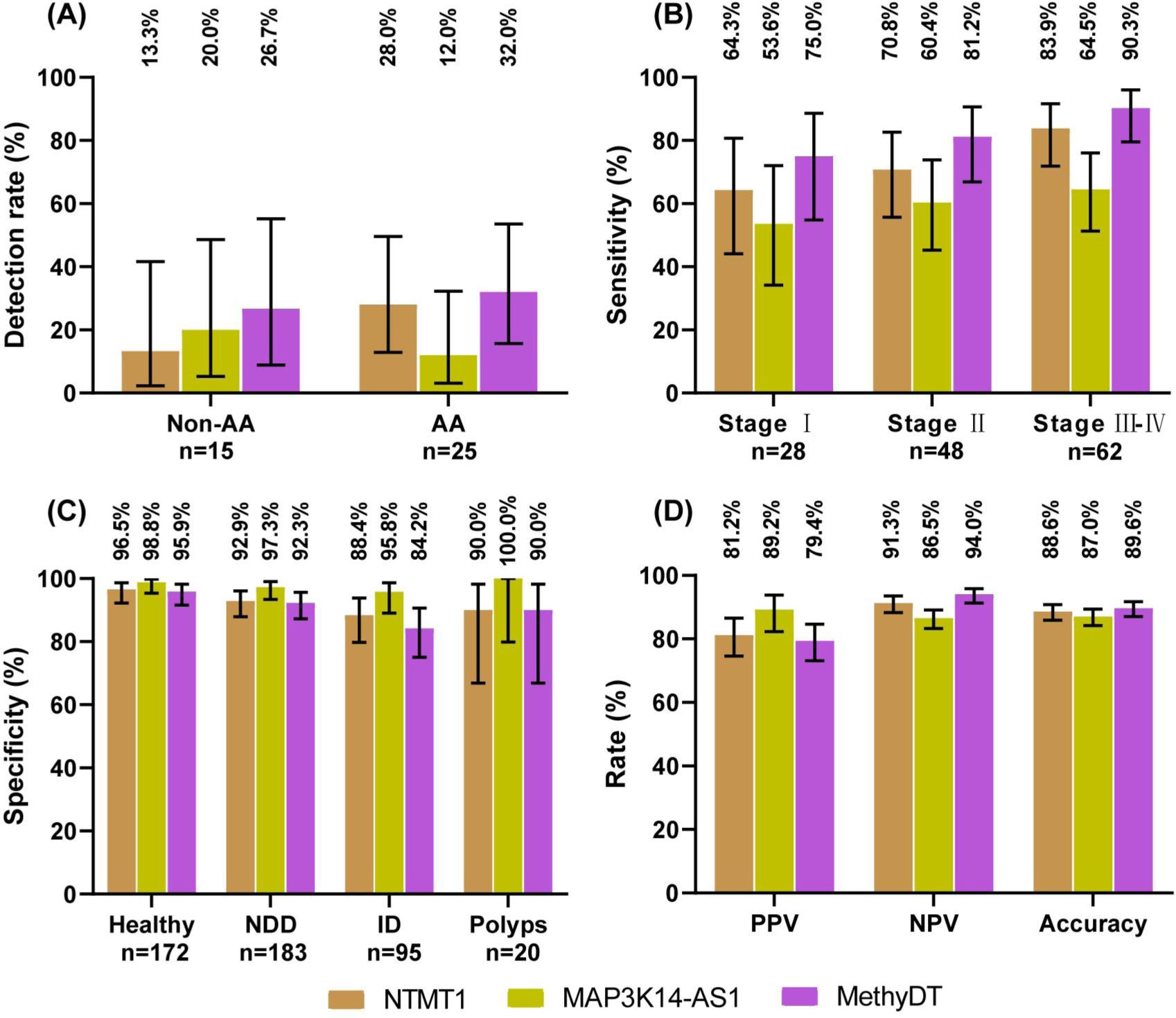
Sensitivity, specificity in various clinical groups and other performance indicators of single biomarker and dual-target in total plasma sample set. (**A**-**B**) Sensitivity in Non-AA and AA groups (A), stages subgroups of CRC (B). (**C**) Specificity in healthy, NDD, ID and polyps groups. (**D**) PPV, NPV and accuracy of individual biomarkers and dual-target in CRC vs control groups. Control groups include healthy, NDD, ID, polyps, Non-AA and other cancers. Error bars represent 95% CI. NDD: Non-digestive disease, ID: Intestinal disease, AA: advanced adenoma.

With regard to the detection rates in adenoma and different stages of CRC, the dual-target test demonstrated superior performance compared to that of a single biomarker (**Figure 7A-B**). With respect to specificities in subgroups, the dual-target test exhibited a marginal decline in value compared to that of a single biomarker (**Figure 7C**). However, with regard to NPV and accuracy, the dual-target test demonstrated superior performance compared to that of a single biomarker (**Figure 7D**). With regard to PPV, the dual-target test exhibited a slight decline in performance relative to that of a single biomarker (**Figure 7D**).

## 4. Discussion

It is commonly accepted that patients diagnosed with CRC at an early stage can be treated more effectively and have a better prognosis. Several stool DNA-based tests have been developed that demonstrate excellent performance in detecting CRCs at their early stages [23-26]. Blood sampling is more acceptable than stool sampling, but blood-based tests are less reported and usually exhibit lower sensitivities than stool-DNA tests, ranging from 47% to 87% [27]. This study presented a systematic pipeline for the discovery of methylation markers from scratch, test development, and evaluation in training and validation plasma sets. The SADMP technique enhanced the ability to detect methylation signals. Following a comprehensive evaluation, the test obtained an overall sensitivity and specificity of 84.8% and 91.5%, respectively, for the detection of CRC at a volume of 2 ml plasma.

Adenoma and CRC displayed lower methylation levels overall, with the exception of regulatory regions (**Figure 2B and 3B**), a finding that has been reported in previous study [17]. Previous studies have focused on the CpG Island Methylator Phenotype (CIMP) found in CRC. This study found that tubular adenomas are common in the methy-L subclass, while villous adenomas are more often in methy-H subclass. Previous studies have indicated that CIMP is rarely found in tubular adenomas, but frequently in tubulovillous and villous adenomas [28], which is in accordance with this study. The large proportion of overlapping DMCs between cancer vs normal and adenoma vs normal indicated that many CpGs have undergone aberrant methylation events at the precancerous stage, which provides robust evidence for discovering the methylation markers for CRC early detection. It should be noted that the CRC markers are not necessarily applicable to adenomas, as a significant proportion of the DMCs in the Cancer vs Normal group were not present in the Adenoma vs Normal group, as illustrated in **Figure 3C**. This emphasises the importance of this study’s inclusion of adenoma samples in the marker discovery set and the selection of DMCs in which cancers and adenomas overlap in a way that many other studies of CRC markers have not done.

Blood-based tests are more susceptible to interfering diseases and may result in a high false-positive rate. Therefore, specificity is a critical indicator. In the marker discovery step, 31 cancer types in the TCGA database and WBC were used to control the low methylation levels of candidate markers in other tissues and blood background, effectively attenuating the interference of unintended cfDNAs. In the assay development phase, we designed highly selective MSP primers that did not show normal amplification curves even when unmethylated DNAs were used as templates at 10^7^ copies (**Figure 5E and 5J**). These aforementioned measures guarantee a high specificity in plasma samples. Two combination algorithms were utilized to assess the performance of the dual-target test in the training set, and both algorithms indicated that the combined markers had better AUC values and higher sensitivities than any single marker (**Figure 6D and 6E**). However, the dual-target test showed a decreased specificity compared to both single markers, from 92.8% and 97.0% to 91.0% (**Figure 6H**), which was also observed in other studies [29,30]. When healthy individuals were selected as controls, the specificity improved to 95.9% (**Figure 7C**), which is comparable to the *Septine9* test [2]. These data demonstrate the excellent performance of the dual-target test in detecting CRC.

A number of studies [31-35] have demonstrated that the methylation levels of *NTMT1* (whose antisense chain counterpart is C9orf50) and *MAP3K14-AS1* can be employed for the screening and diagnosis of CRC. In particular, the study conducted by Sarah Ø Jensen et al [34] indicated that the C9orf50 methylation assay exhibited a plasma sensitivity of 76% and specificity of 91% for CRC. Ludovic Barault and colleagues [35] demonstrated that the *MAP3K14-AS1* methylation assay in plasma exhibited a sensitivity of 69.8% and a specificity of 100% for CRC. The dual-strand technique has been proven to enhance the performance of markers in previous studies [14,15]. This technique was also observed to be effective in our study. By detecting the methylation signals of *NTMT1* sense- and antisense-strand simultaneously, the Ct value of the dual-strand assay was able to shift forward by one compared to the single-strand assay (**Figure 5D**). In contrast to previous studies, the current study also included two MGB probes located downstream of forward and reverse primers of *MAP3K14-AS1*. During PCR strand extension, the polymerase enzymes cleaved the 5-primer sequence of probes and released two fluorescent groups. The dual-MGB probe technique doubled the fluorescent signals when both probes shared the same channel, leading to an earlier Ct value similar to that of the dual-strand technique (**Figure 5I**). Serial dilution experiments confirmed the superiority of dual-MGB probes over one MGB probe (**Figure 5H-J**). These results suggest that applying the SADMP technique can be a feasible strategy to enhance the detection sensitivity of candidate markers.

Early diagnosis or screening techniques are essential to improve patient survival time, particularly when curable treatments are available. Studies have shown that the 5-year survival rate of early detected CRC is almost 90%, while it was only 20% for advanced CRC [36]. The dual-target test showed a sensitivity of 75.0% and 81.2% for stage I and stage II CRC detection, notably, the dual-target test obtained a positive detection rate of 32.00% (8/25) for AA (**Figure 7B and 7C**), implying its ability to detect early CRC and precancerous lesions.

The current study has some limitations that may hamper the interpretation of these results. 1) Participants in this study were enrolled from a single center, which may bias these results. 2) the SADMP techniques may not applicable for all candidate markers. The dual-strand technique may be attempted when both sense and antisense strands are suitable for designing MSP primers, while the multiple MGB probe technique is limited by the amplicon length, which is usually less than 100 bp.

## 5. Conclusions

In this study, we employed several public databases of adenomas and CRC for marker discovering, and ultimately identified two promising markers, *NTMT1* and *MAP3K14-AS1*. We then constructed the SADMP technology based on these two markers, which enhanced the sensitivity of the detection. The dual-target assay has a high sensitivity for AA and early stage CRC, and its clinical application value merits further investigation.

## Supplementary Materials

The following supporting information can be downloaded at: www.mdpi.com/xxx/s1, Figure S1: The methylation status of candidate markers verified by Sanger sequencing; Table S1: The clinical features of training and validation cohorts used in this study; Table S2: The information of primers for Sanger sequencing in this study; Table S3: The information of primers and MGB probes used in this study; Table S4: The amplification system of the MSP.

## Author Contributions

Conception: YT Z, ZJ W and SC W; Supervision: YT Z and SC W; Funding acquisition: YT Z; Project administration: ZJ W; Methodology, Resources, Data curation: QN Y, X L, X L, SL D, XP L and T Z; Sofeware, Validation, Formal analysis: DH Z; Writing-original draft: YT Z; Writing-review & editing: YT Z, ZJ W and SC W. All authors have read and agreed to the published version of the manuscript.

## Institutional Review Board Statement

The study was conducted in accordance with the Declaration of Helsinki, and approved by the Ethics Committee of the First Affiliated Hospital of Zhengzhou University (approval number: 2022-KY-0631-002).

## Informed Consent Statement

Informed consent was obtained from all subjects involved in the study.

## Data Availability Statement

The datasets (GSE77954, GSE101764, GSE131013, GSE164811, GSE193535, GSE129364, GSE139404, GSE107352, GSE75546, GSE77965, GSE199057, GSE68060, GSE48684, GSE40279 and GSE122126) supporting the conclusions of this article are available in the Gene Expression Omnibus database (https://www.ncbi.nlm.nih.gov/geo/query/acc.cgi). The E-MTAB-6450 data is available in EMBL biostudies (https://www.ebi.ac.uk/biostudies/arrayexpress). The source code and intermediate data can be accessed from git-hub (https://github.com/amsinfor/CRC-methylation).

## Acknowledgments

We thank Dr. Kangkang wan for the assistance in MSP designing and data analysis in this work.

---

**Supplemental table 1.** The clinical features of training and validation cohorts used in this study.

|  | Training set |  |  |  |  |  |  |  | Validation set |  |  |  |  |  |  |  |
| --- | --- | --- | --- | --- | --- | --- | --- | --- | --- | --- | --- | --- | --- | --- | --- | --- |
|  | Healthy<br>n=115 | NDD<br>n=123 | ID<br>n=65 | Polyps<br>n=14 | Non-AA<br>n=10 | AA<br>n=16 | CRC<br>n=125 | Other<br>cancers<br>n=6 | Healthy<br>n=57 | NDD<br>n=60 | ID<br>n=30 | Polyps<br>n=6 | Non-AA<br>n=5 | AA<br>n=9 | CRC<br>n=66 | Other<br>cancers<br>n=2 |
| Sex (n, %) |  |  |  |  |  |  |  |  |  |  |  |  |  |  |  |  |
| Male | 65<br>(56.5%) | 63<br>(51.2%) | 38<br>(58.5%) | 10<br>(71.4%) | 7<br>(70.0%) | 7<br>(43.8%) | 70<br>(56.0%) | 4<br>(66.7%) | 25<br>(43.9%) | 39<br>(65.0%) | 12<br>(40.0%) | 3<br>(50.0%) | 3 (60.0%) | 4<br>(44.4%) | 37<br>(56.1%) | 1<br>(50.0%) |
| Female | 50<br>(43.5%) | 60<br>(48.8%) | 27<br>(41.5%) | 4<br>(28.6%) | 3<br>(30.0%) | 9<br>(56.3%) | 55<br>(44.0%) | 2<br>(33.3%) | 32<br>(56.1%) | 21<br>(35.0%) | 18<br>(60.0%) | 3<br>(50.0%) | 2 (40.0%) | 5<br>(55.6%) | 29<br>(43.9%) | 1<br>(50.0%) |
| Age |  |  |  |  |  |  |  |  |  |  |  |  |  |  |  |  |
| Median | 40 | 52 | 62 | 66 | 65 | 56 | 64 | 65 | 40 | 50 | 58 | 64 | 53 | 54 | 64 | 69 |
| Range | 25-45 | 33-80 | 33-79 | 39-83 | 52-73 | 43-76 | 33-86 | 41-80 | 25-43 | 30-60 | 40-80 | 44-74 | 49-67 | 38-76 | 33-82 | 59-79 |
| TNM Stage<br>(n, %) |  |  |  |  |  |  |  |  |  |  |  |  |  |  |  |  |
| I |  |  |  |  |  |  | 20<br>(16.0%) |  |  |  |  |  |  |  | 8 (12.1%) |  |
| II |  |  |  |  |  |  | 33<br>(26.4%) |  |  |  |  |  |  |  | 15<br>(22.7%) |  |
| III |  |  |  |  |  |  | 37<br>(29.6%) |  |  |  |  |  |  |  | 23<br>(34.8%) |  |
| IV |  |  |  |  |  |  | 2 (1.6%) |  |  |  |  |  |  |  | 0 (0.0%) |  |
| Na |  |  |  |  |  |  | 33<br>(26.4%) |  |  |  |  |  |  |  | 20<br>(30.3%) |  |

**Supplemental Table 2.**
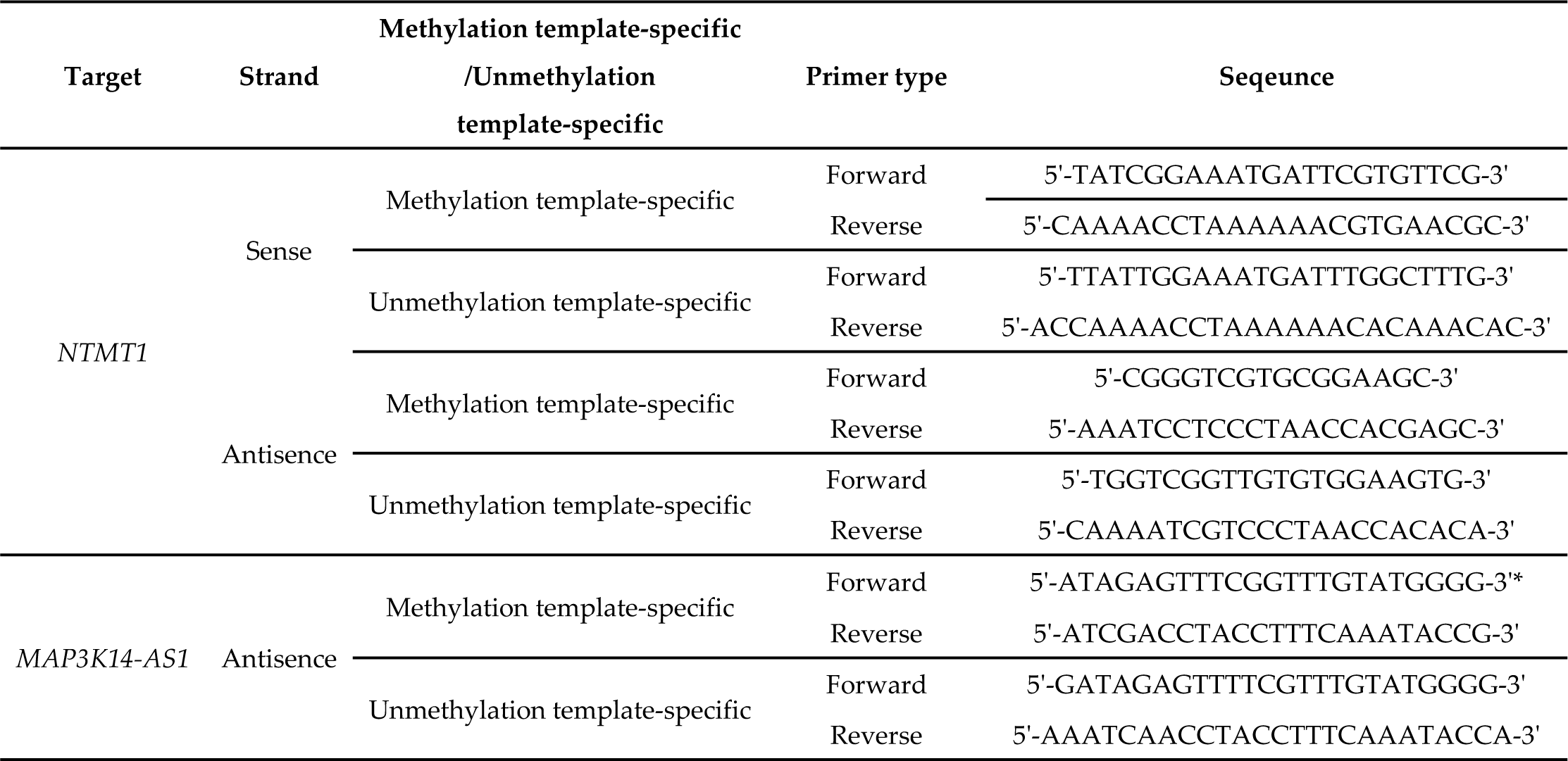
The information of primers for Sanger sequencing in this study.

**Supplemental Table 3.**
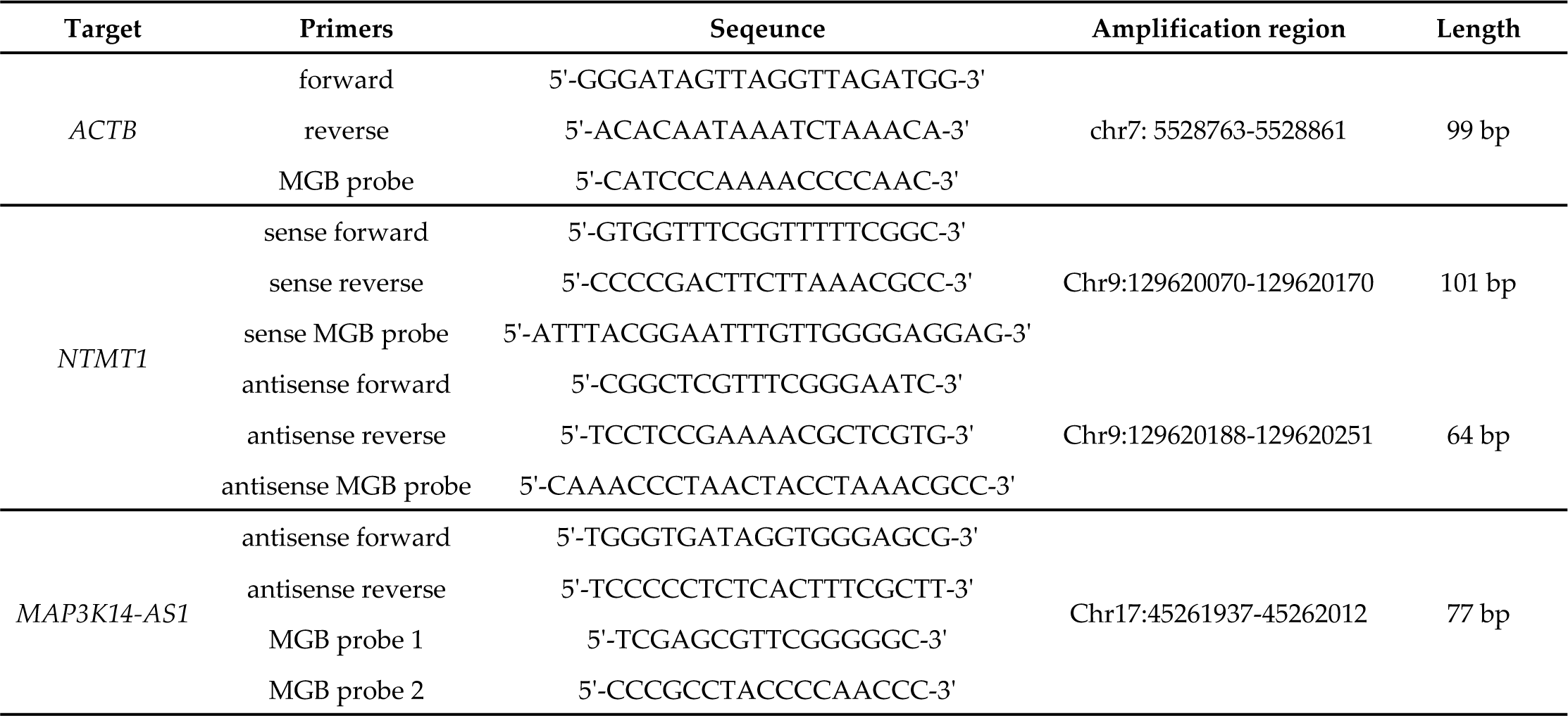
The information of primers and MGB probes used in this study.

**Supplemental Table 4.**
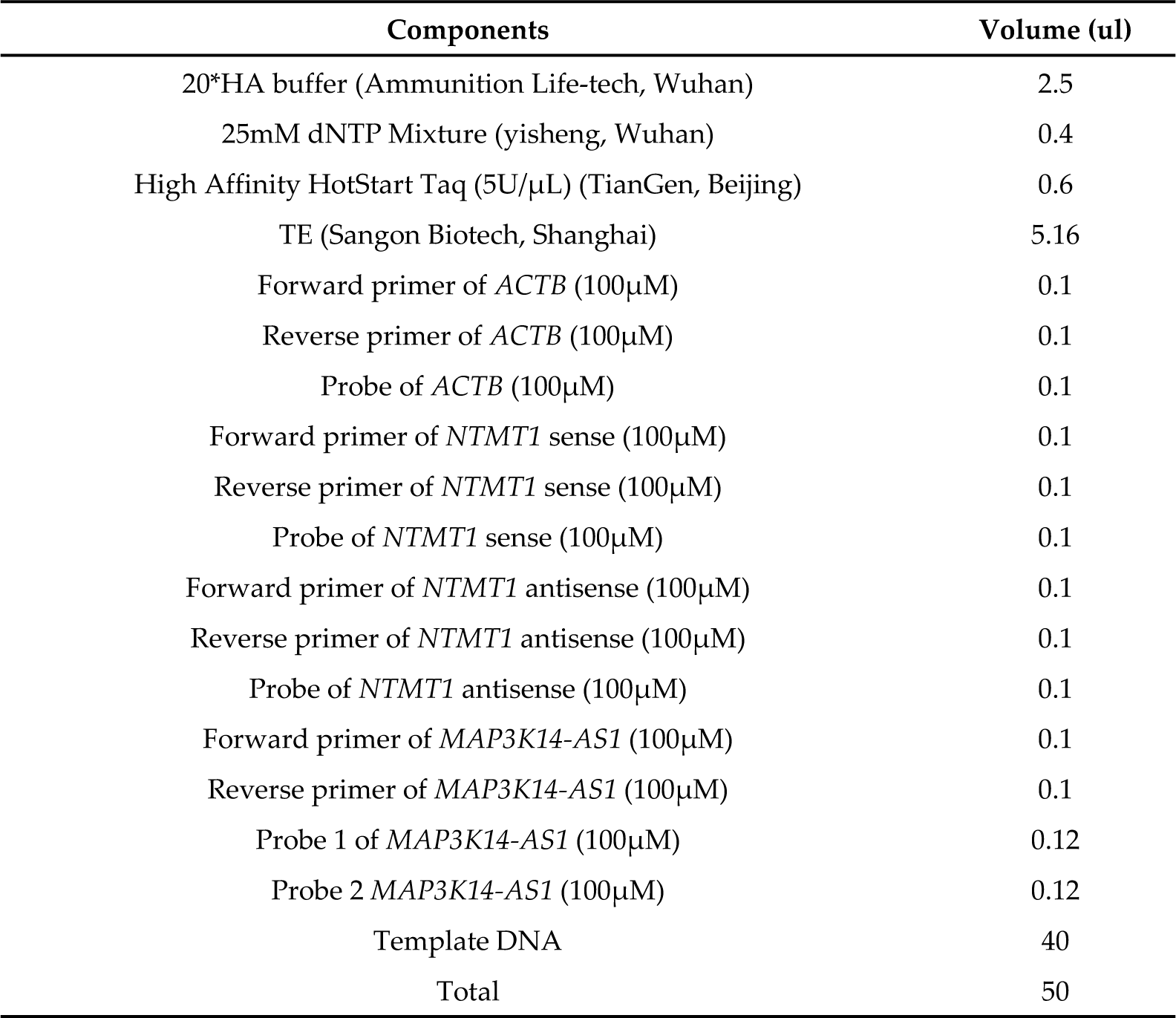
The amplification system of the MSP.

## Disclaimer/Publisher’s Note

The statements, opinions and data contained in all publications are solely those of the individual author(s) and contributor(s) and not of MDPI and/or the editor(s). MDPI and/or the editor(s) disclaim responsibility for any injury to people or property resulting from any ideas, methods, instructions or products referred to in the content.

## References

1. Powrózek T, Krawczyk P, Kucharczyk T, Milanowski J. Septin 9 promoter region methylation in free circulating dna—potential role in noninvasive diagnosis of lung cancer: preliminary report. Med Oncol 2014;31: 917. doi: 10.1007/s12032-014-0917-4

2. Lamb YN, Dhillon S. Epi procolon® 2.0 ce: a blood-based screening test for colorectal cancer. Mol Diagn Ther 2017;21: 225–32. doi: 10.1007/s40291-017-0259-y

3. Sun J, Fei F, Zhang M, Li Y, Zhang X, Zhu S, et al. The role of msept9 in screening, diagnosis, and recurrence monitoring of colorectal cancer. Bmc Cancer 2019; 19: 450. doi: 10.1186/s12885-019-5663-8

4. Jung G, Hernández-Illán E, Moreira L, Balaguer F, Goel A. Epigenetics of colorectal cancer: biomarker and therapeutic potential. Nat Rev Gastroenterol Hepatol 2020;17(2):111–130. doi: 10.1038/s41575-019-0230-y.

5. Church TR, Wandell M, Lofton-Day C, Mongin SJ, Burger M, Payne SR, et al. Prospective evaluation of methylated SEPT9 in plasma for detection of asymptomatic colorectal cancer. Gut 2014; 63(2):317–25. doi: 10.1136/gutjnl-2012-304149.

6. Nassar FJ, Msheik ZS, Nasr RR, Temraz SN. Methylated circulating tumor DNA as a biomarker for colorectal cancer diagnosis, prognosis, and prediction. Clin Epigenetics 2021;13(1):111. doi: 10.1186/s13148-021-01095-5.

7. Li B, Huang H, Huang R, Zhang W, Zhou G, Wu Z, et al. Sept9 gene methylation as a noninvasive marker for hepatocellular carcinoma. Dis Markers 2020; 2020: 6289063. doi: 10.1155/2020/6289063

8. Cao CQ, Chang L, Wu Q. Circulating methylated septin 9 and ring finger protein 180 for noninvasive diagnosis of early gastric cancer. Transl Cancer Res 2020; 9: 7012–21. doi: 10.21037/tcr-20-1330

9. Jiao X, Zhang S, Jiao J, Zhang T, Qu W, Muloye GM, et al. Promoter methylation of sept9 as a potential biomarker for early detection of cervical cancer and its overexpression predicts radioresistance. Clin Epigenetics 2019;11: 120. doi: 10.1186/s13148-019-0719-9

10. Matsui S, Kagara N, Mishima C, Naoi Y, Shimoda M, Shimomura A, et al. Methylation of the sept9_v2 promoter as a novel marker for the detection of circulating tumor dna in breast cancer patients. Oncol Rep 2016; 36: 2225–35. doi: 10.3892/or.2016.5004

11. Song P, Wu LR, Yan YH, Zhang JX, Chu T, Kwong LN, et al. Limitations and opportunities of technologies for the analysis of cell-free DNA in cancer diagnostics. Nat Biomed Eng 2022;6(3):232–245. doi: 10.1038/s41551-021-00837-3.

12. Babayan A, Pantel K. Advances in liquid biopsy approaches for early detection and monitoring of cancer. Genome Med 2018; 10: 21. doi: 10.1186/s13073-018-0533-6

13. Li LC, Dahiya R. Methprimer: designing primers for methylation pcrs. Bioinformatics 2002;18: 1427–31. doi: 10.1093/bioinformatics/18.11.1427

14. Jensen SØ, Øgaard N, Nielsen HJ, Bramsen JB, Andersen CL. Enhanced performance of dna methylation markers by simultaneous measurement of sense and antisense dna strands after cytosine conversion. Clin Chem 2020; 66: 925–33. doi: 10.1093/clinchem/hvaa100

15. Faaborg L, Fredslund AR, Waldstrøm M, Høgdall E, Høgdall C, Adimi P, et al. Analysis of hoxa9 methylated ctdna in ovarian cancer using sense-antisense measurement. Clin Chim Acta 2021; 522: 152–7. doi: 10.1016/j.cca.2021.08.020

16. Aryee MJ, Jaffe AE, Corrada-Bravo H, Ladd-Acosta C, Feinberg AP, Hansen KD, et al. Minfi: a flexible and comprehensive bioconductor package for the analysis of infinium dna methylation microarrays. Bioinformatics 2014; 30: 1363–9. doi: 10.1093/bioinformatics/btu049

17. Luo Y, Wong CJ, Kaz AM, Dzieciatkowski S, Carter KT, Morris SM, et al. Differences in dna methylation signatures reveal multiple pathways of progression from adenoma to colorectal cancer. Gastroenterology 2014; 147: 418–29. doi: 10.1053/j.gastro.2014.04.039

18. Hannum G, Guinney J, Zhao L, Zhang L, Hughes G, Sadda S, et al. Genome-wide methylation profiles reveal quantitative views of human aging rates. Mol Cell 2013; 49: 359–67. doi: 10.1016/j.molcel.2012.10.016

19. Moss J, Magenheim J, Neiman D, Zemmour H, Loyfer N, Korach A, et al. Comprehensive human cell-type methylation atlas reveals origins of circulating cell-free dna in health and disease. Nat Commun 2018; 9: 5068. doi: 10.1038/s41467-018-07466-6

20. Li R, Qu B, Wan K, Lu C, Li T, Zhou F, et al. Identification of two methylated fragments of an sdc2 cpg island using a sliding window technique for early detection of colorectal cancer. Febs Open Bio. (2021) 11: 1941–52. doi: 10.1002/2211-5463.13180

21. Zhang L, Dong L, Lu C, Huang W, Yang C, Wang Q, et al. Methylation of SDC2/TFPI2 and Its Diagnostic Value in Colorectal Tumorous Lesions. Front Mol Biosci. 2021;8:706754. doi: 10.3389/fmolb.2021.706754.

22. Bian Y, Gao Y, Lu C, Tian B, Xin L, Lin H, et al. Genome-wide methylation profiling identified methylated KCNA3 and OTOP2 as promising diagnostic markers for esophageal squamous cell carcinoma. Chin Med J (Engl) 2023; doi: 10.1097/CM9.0000000000002832.

23. Wang Z, Shang J, Zhang G, Kong L, Zhang F, Guo Y, et al. Evaluating the clinical performance of a dual-target stool dna test for colorectal cancer detection. J Mol Diagn 2022; 24: 131–43. doi: 10.1016/j.jmoldx.2021.10.012

24. Imperiale TF, Ransohoff DF, Itzkowitz SH, Levin TR, Lavin P, Lidgard GP, et al. Multitarget stool dna testing for colorectal-cancer screening. N Engl J Med 2014;370: 1287–97. doi: 10.1056/NEJMoa1311194

25. Zhao G, Liu X, Liu Y, Li H, Ma Y, Li S, et al. Aberrant dna methylation of sept9 and sdc2 in stool specimens as an integrated biomarker for colorectal cancer early detection. Front Genet 2020; 11: 643. doi: 10.3389/fgene.2020.00643

26. Oh TJ, Oh HI, Seo YY, Jeong D, Kim C, Kang HW, et al. Feasibility of quantifying sdc2methylation in stool dna for early detection of colorectal cancer. Clin Epigenetics 2017; 9: 126. doi: 10.1186/s13148-017-0426-3

27. Nassar FJ, Msheik ZS, Nasr RR, Temraz SN. Methylated circulating tumor dna as a biomarker for colorectal cancer diagnosis, prognosis, and prediction. Clin Epigenetics 2021; 13: 111. doi: 10.1186/s13148-021-01095-5

28. Kakar S, Deng G, Cun L, Sahai V, Kim YS. Cpg island methylation is frequently present in tubulovillous and villous adenomas and correlates with size, site, and villous component. Hum Pathol 2008; 39: 30–6. doi: 10.1016/j.humpath.2007.06.002

29. Zhao G, Li H, Yang Z, Wang Z, Xu M, Xiong S, et al. Multiplex methylated dna testing in plasma with high sensitivity and specificity for colorectal cancer screening. Cancer Med 2019; 8: 5619–28. doi: 10.1002/cam4.2475

30. Bagheri H, Mosallaei M, Bagherpour B, Khosravi S, Salehi AR, Salehi R. Tfpi2 and ndrg4 gene promoter methylation analysis in peripheral blood mononuclear cells are novel epigenetic noninvasive biomarkers for colorectal cancer diagnosis. J Gene Med 2020;22: e3189. doi: 10.1002/jgm.3189

31. Cao Y, Zhao G, Yuan M, Liu X, Ma Y, Cao Y, et al. KCNQ5 and C9orf50 Methylation in Stool DNA for Early Detection of Colorectal Cancer. Front Oncol 2021; 29;10:621295. doi: 10.3389/fonc.2020.621295.

32. Zhang Y, Wu Q, Xu L, Wang H, Liu X, Li S, et al. Sensitive detection of colorectal cancer in peripheral blood by a novel methylation assay. Clin Epigenetics 2021;13(1):90. doi: 10.1186/s13148-021-01076-8.

33. Jensen SØ, Øgaard N, Ørntoft MW, Rasmussen MH, Bramsen JB, Kristensen H, et al. Novel DNA methylation biomarkers show high sensitivity and specificity for blood-based detection of colorectal cancer-a clinical biomarker discovery and validation study. Clin Epigenetics 2019;11(1):158. doi: 10.1186/s13148-019-0757-3.

34. Barault L, Amatu A, Siravegna G, Ponzetti A, Moran S, Cassingena A, et al. Discovery of methylated circulating DNA biomarkers for comprehensive non-invasive monitoring of treatment response in metastatic colorectal cancer. Gut 2018; 67(11):1995–2005. doi: 10.1136/gutjnl-2016-313372.

35. Huang H, Cao W, Long Z, Kuang L, Li X, Feng Y, et al. DNA methylation-based patterns for early diagnostic prediction and prognostic evaluation in colorectal cancer patients with high tumor mutation burden. Front Oncol 2023;12:1030335. doi: 10.3389/fonc.2022.1030335.

36. Ladabaum U, Dominitz JA, Kahi C, Schoen RE. Strategies for colorectal cancer screening. Gastroenterology 2020; 158: 418–32. doi: 10.1053/j.gastro.2019.06.043

